# Impact of vaccination with SCB-2019 COVID-19 vaccine on transmission of SARS-CoV-2 infection: a household contact study in the Philippines

**DOI:** 10.1101/2022.08.18.22278764

**Authors:** Birkneh Tilahun Tadesse, Lulu Bravo, Florian Marks, Asma Binte Aziz, Young Ae You, Jonathan Sugimoto, Ping Li, Joyce Garcia, Frank Rockhold, Ralf Clemens, the HHC study group

## Abstract

**Background:** An exploratory household transmission study was nested in SPECTRA, the phase 2/3 efficacy study of the adjuvanted recombinant protein-based COVID-19 vaccine SCB-2019. We compared occurrence of confirmed COVID-19 infections between households and household contacts of infected SPECTRA participants who were either placebo or SCB-2019 recipients.

**Methods:** SPECTRA trial participants at eight study sites in the Philippines who developed rRT-PCR-confirmed COVID-19 were contacted by a study team blinded to assignment of index cases to vaccine or placebo groups to enroll in this household transmission study. Enrolled households and household contacts were monitored for three weeks using rRT-PCR and rapid antigen testing to detect new COVID-19 infections.

**Results:** Observation of the households of 154 eligible COVID-19 index cases, 130 symptomatic and 24 asymptomatic at diagnosis, revealed household secondary attack rates for any COVID-19 infection of SCB-2019 index cases of 0.76% (90% CI: 0.15–3.90) compared with 5.88% (90% CI: 3.20–10.8) in placebo index case households, a relative risk reduction of 79% (90% CI: -28–97). The relative risk reduction of symptomatic COVID-19 was 84% (90% CI: 28–97) for household contacts of all COVID-19 infected index cases, and 80% (90% CI: 7–96) for household contacts of index cases with symptomatic COVID-19.

**Conclusions:** In this prospective household contact study vaccination with SCB-2019 reduced SARS-CoV-2 transmission in households, so decreasing infections of household contacts, compared with placebo.

## INTRODUCTION

Almost 550 million people have contracted COVID-19 due to infection by the severe acute respiratory syndrome-related coronavirus (SARS-CoV-2) [1] with more than 18 million excess deaths worldwide [2]. Transmission is through three main mechanisms: inhalation, direct inoculation onto mucosal surfaces, or touching contaminated surfaces [3]. Airborne transmission has been argued to be the dominant mechanism [4,5], infected individuals releasing virus in droplets during coughing, sneezing, talking, or singing to infect others [6], leading to the global implementation of mask-wearing policies in public places.

As risk of infection directly relates to duration and proximity to an index case their household members are estimated to be at a tenfold higher risk of infection [7,8]. Overall secondary attack rates (SAR) in household contacts (HHC) have been estimated to be 3% to 40% [9–11], with highest SAR reported following symptomatic index cases (18%), spouse contacts (37.8%) and in households with only one contact (41%) [11].

Globally, after masking and social distancing measures, the main COVID-19 prevention strategy has been mass immunization to provide personal protection and minimize community and household transmission. Therefore, impact of any new SARS-CoV-2 vaccine on preventing household transmission is an essential aspect of vaccine development. There is limited evidence on vaccines preventing SARS-CoV-2 infection of HHC although several investigations have reported positive impact [12–18]. A database link study reported 50% reduction among HHC of vaccinated versus unvaccinated index cases [13], but other studies found limited impact of vaccination especially for index cases with the Delta (B.1.617.2) variant [17,18], potentially due to the high viral load in breakthrough cases. We report a prospective exploratory clinical study of the impact of vaccination with SCB-2019, an adjuvanted recombinant protein-based COVID-19 vaccine, on household transmission in households of index cases who previously received SCB-2019 or placebo.

## METHODS

This exploratory study was conducted alongside the pivotal, randomized, placebo-controlled, phase 2/3 efficacy trial of SCB-2019 vaccine (SPECTRA; ClinicalTrials.gov NCT04672395) [19]. The objective of this household transmission (HHT) investigation, performed in eight of the SPECTRA Philippine sites, was to assess and compare reductions in COVID-19 infections in households and household members of SCB-2019 vaccine recipients with a breakthrough infection with those in households of placebo recipients. Sites and numbers of placebo or vaccine recipient index cases enrolled by site are presented in *Supplementary Table 1*. The protocol was approved by the Single Joint Research Ethics Board (SJREB), the respective Institutional Review Boards (IRB) of the eight study sites and the International Vaccine Institute IRB and implemented in accordance with the Declaration of Helsinki, principles of ICH-GCP and the Philippines’ ethical requirements. All participants or their parents or legal guardians provided written informed consent or assent.

### Participants and procedures

Participants of SPECTRA were followed as part of the main trial using symptom screening and weekly rapid antigen testing (RAT) for COVID-19 infection [19]. Any participant at one of the eight HHT study sites who developed COVID-19 confirmed by a positive real-time reverse transcriptase polymerase chain reaction (rRT-PCR) during follow-up was contacted by the HHT study team for their willingness to enroll in this HHT study and for follow-up of their household. The HHT study team was blinded to the randomization of the index cases to vaccine or placebo groups in the SPECTRA trial. Once the trial participant consented to participate in the HHT study their HHC were contacted for their consent within 3–5 days for inclusion and three weeks of follow-up in the HHT study. Eligible HHC were 6 years-old or older, with no other household member other than the participant index case to have been co-diagnosed with COVID-19.

Key sociodemographic and vaccination information including age, sex, income status, COVID-19 risk and COVID-19 preventive efforts (masking, crowd avoidance, and handwashing/sanitizer use), and presence of any comorbidities was obtained by interview of enrolled participants. Solicited COVID-19 information included any suggestive symptoms (breathing difficulties, fever, chills, cough, fatigue, muscle/body aches, headache, new loss of taste or smell, sore throat, congestion or runny nose, nausea, vomiting, or diarrhea), and vaccination history included vaccine type, date of vaccination, complete or partial vaccination. All consented HHC without COVID-19 symptoms were tested at enrollment for anti-SARS-CoV-2 IgM or IgG/IgM to exclude any acute asymptomatic infections. Nasopharyngeal (NP) swab samples were taken for RT-PCR testing from any HHC who reported symptoms suggestive of COVID-19 or had a positive antibody test.

### Follow-up for COVID-19 symptoms and NP sampling

Enrolled HHC completed daily symptom checklists for any COVID-19 symptoms for three weeks which were reviewed by study coordinators available 24 hours a day, 7 days a week. HHC who developed any symptoms were instructed to promptly contact the study coordinator and NP samples collected at their home or the study site were transported to one of two certified molecular laboratories in the Philippines – Manila Doctors Hospital or the Asian Hospital and Medical Center to detect SARS-CoV-2 by rRT-PCR using the manufacturer protocols, with molecular assays and specific laboratory instructions according to WHO guidelines [20]. Study coordinators telephoned households weekly to review the symptom checklist for any symptoms that had been missed. In cases of unreported mild symptoms, NP samples were collected on the day of identification.

rRT-PCR-confirmed cases were considered as symptomatic COVID-19 and were followed for severity from time of diagnosis until completion of treatment or outcome 14 days after symptom onset. For symptomatic COVID-19 infections in HHC a detailed case report was completed to capture risk of infection, relationship to index case, any contact with known or suspected COVID-19 cases other than the index case within the previous 2 weeks, and details of symptoms including temporal relationship with the index case diagnosis.

Asymptomatic SARS-CoV-2 infections during the three-week follow-up were determined using lateral flow anti-N antibody rapid antibody test (RAT) kits (PCL, South Korea) [21]. Household participants with negative antibody tests at baseline when the index case was diagnosed, and no symptoms or positive test during the surveillance, were retested at the end of follow-up. Those with a positive IgM or IgG/IgM provided an NP sample for rRT-PCR while those with IgG positivity were followed for any symptoms during the follow-up. Similarly, IgM positivity was considered as a sign of acute infection and an indication for PCR testing. All HHC had an exit follow-up visit at the study sites.

### Statistical Analysis

Baseline characteristics (COVID-19 risk, co-morbidities and COVID-19 vaccination status) of household contacts of SCB-2019 and placebo index cases were summarized and compared using Chi-square or Fisher’s exact test for categorical variables, and Student’s t-test, or Wilcoxon rank sum test for or non-parametric tests depending on whether the data were normally distributed.

Primary endpoints were the symptomatic secondary attack rates (SAR) in households of SCB-2019 and placebo recipients in the SPECTRA trial (index cases), secondary endpoints were asymptomatic COVID-19 infections in those households. Point estimates and 90% confidence intervals of symptomatic, asymptomatic, and overall SAR by household and the relative risk reduction (RRR) were estimated using the generalized estimating equation (GEE) logistic regression modeling approach with jackknifed standard errors, using the geeglm function of the geepack library of R (version 4.1.2) and an exchangeable correlation structure. The percentage of HHC vaccinated was used for SAR estimation stratified by COVID-19 vaccination. For SARs by household, the estimates were adjusted for vaccination coverage in the households, which was calculated as the proportion of HHC who received the vaccine in a single household. Vaccination coverage in contacts in each household were categorized in to 0–24%, 24–49%, 50–74% and 75–100% for inclusion in the models. For the SAR estimation by HHC, however, the proportion of HHC who were fully or partially vaccinated or were unvaccinated in each treatment group of the index cases were used to adjust the SAR estimation.

## RESULTS

During active surveillance for this HHT study from July 9, 2021 to December 6, 2021 we identified 598 rRT-PCR-confirmed index cases among SPECTRA trial participants of whom 117 said they had no eligible household members and 42 were detected too late (more than 5 days after diagnosis of the index case); 184 of the remaining 439 did not consent to having their household members contacted and 98 households refused to participate. Of the 157 index cases enrolled two were excluded as they lived in the same household and a further case was found to have no eligible household member. Thus, 154 index cases representing 154 households; 51 SCB-2019 and 103 placebo index cases, respectively, were eligible (**Figure 1**), 130 (42 SCB-2019 and 88 placebo) symptomatic and 24 (9 SCB-2019 and 15 placebo) asymptomatic at diagnosis. Sequencing analysis of 74 index case swab samples, 24 in SCB-2019 and 50 in placebo groups, identified the Delta (B.1.617.2) variant in all cases (**Table 1**). Mean ages of index cases were similar in SCB-2019 index cases (36.7 ± 12.2 years) and in placebo groups (35.0 ± 10.9 years) as they were in terms of sex, symptoms, and adherence to the SPECTRA study vaccination regimen (**Table 1**).

**Table 1:**
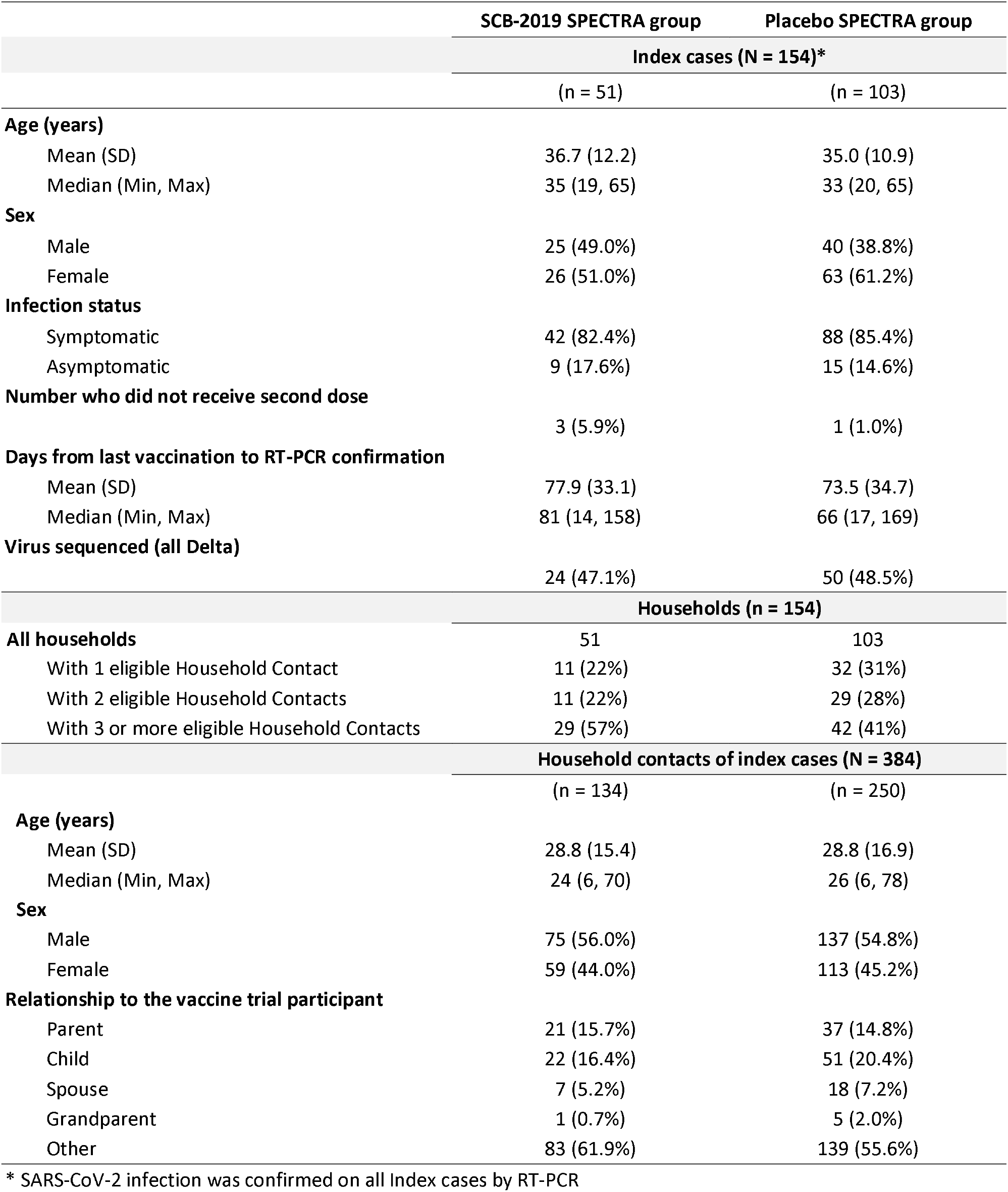
Sociodemographic information of the Index cases and their household contacts

|  | SCB-2019 SPECTRA group | Placebo SPECTRA group |
| --- | --- | --- |
|  | Index cases (N = 154)* |  |
|  | (n = 51) | (n = 103) |
| **Age (years)** |  |  |
| Mean (SD) | 36.7 (12.2) | 35.0 (10.9) |
| Median (Min, Max) | 35 (19, 65) | 33 (20, 65) |
| **Sex** |  |  |
| Male | 25 (49.0%) | 40 (38.8%) |
| Female | 26 (51.0%) | 63 (61.2%) |
| **Infection status** |  |  |
| Symptomatic | 42 (82.4%) | 88 (85.4%) |
| Asymptomatic | 9 (17.6%) | 15 (14.6%) |
| **Number who did not receive second dose** |  |  |
|  | 3 (5.9%) | 1 (1.0%) |
| **Days from last vaccination to RT-PCR confirmation** |  |  |
| Mean (SD) | 77.9 (33.1) | 73.5 (34.7) |
| Median (Min, Max) | 81 (14, 158) | 66 (17, 169) |
| **Virus sequenced (all Delta)** |  |  |
|  | 24 (47.1%) | 50 (48.5%) |
|  | Households (n = 154) |  |
| **All households** | 51 | 103 |
| With 1 eligible Household Contact | 11 (22%) | 32 (31%) |
| With 2 eligible Household Contacts | 11 (22%) | 29 (28%) |
| With 3 or more eligible Household Contacts | 29 (57%) | 42 (41%) |
|  | Household contacts of index cases (N = 384) |  |
|  | (n = 134) | (n = 250) |
| **Age (years)** |  |  |
| Mean (SD) | 28.8 (15.4) | 28.8 (16.9) |
| Median (Min, Max) | 24 (6, 70) | 26 (6, 78) |
| **Sex** |  |  |
| Male | 75 (56.0%) | 137 (54.8%) |
| Female | 59 (44.0%) | 113 (45.2%) |
| **Relationship to the vaccine trial participant** |  |  |
| Parent | 21 (15.7%) | 37 (14.8%) |
| Child | 22 (16.4%) | 51 (20.4%) |
| Spouse | 7 (5.2%) | 18 (7.2%) |
| Grandparent | 1 (0.7%) | 5 (2.0%) |
| Other | 83 (61.9%) | 139 (55.6%) |

\* SARS-CoV-2 infection was confirmed on all Index cases by RT-PCR

**Figure 1.**
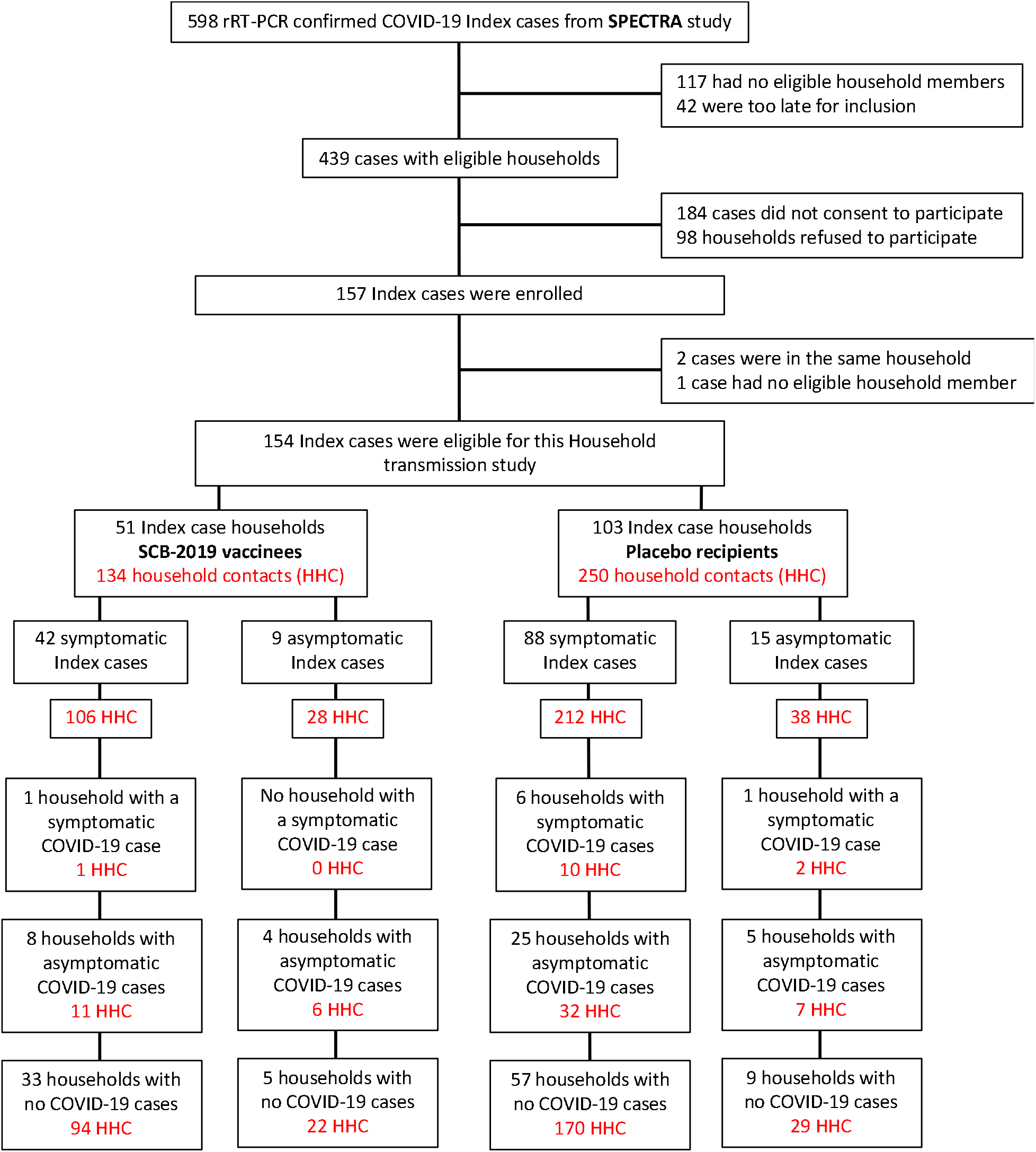
Disposition of households, household contacts (HHC) and household infections by index case SPECTRA treatment group.

A total of 388 HHC agreed to enroll of whom 4 were lost to follow-up with 384 completing the study, 134 HHC of SPECTRA SCB-2019 vaccinees, and 250 HHC of SPECTRA placebo recipients index cases (**Table 1**). As with the index cases, households and HHC had comparable household composition and size, demographic characteristics of age, sex, and relationship to the index case. Vaccination status of HHC (***Supplementary Table 2***) shows that the majority were either not vaccinated or only partially vaccinated, only 22.9% being reported as fully vaccinated, a proportion that was similar in SCB-2019 and placebo groups. Baseline and endline serostatus of HHC as an indication of past or present infection status is presented in **Table 2**. Most infected HHCs were from symptomatic index cases; 106 (79.1%) HHC of 42 (82.4%) index SCB-2019 cases and 212 (84.8%) HHC of 88 (85.4%) placebo recipient symptomatic index cases. The remainder in both groups were contacts of asymptomatic index cases (**Figure 1**).

**Table 2.** COVID-19 infection history and vaccine-related information in household contacts

|  | Household contacts (HHC) |  |
| --- | --- | --- |
|  | SCB-2019<br>(n = 134) | Placebo<br>(n = 250) |
| **COVID-19 tested in the last 3 months, n (%)** |  |  |
| All tested | 12 (9.0%) | 29 (11.6%) |
| Positive test result | 1 (0.7%) | 3 (1.2%) |
| Negative test result | 11 (8.2%) | 26 (10.4%) |
| **Vaccination status for COVID-19, n (%)** |  |  |
| Unvaccinated | 85 (63.4%) | 154 (61.6%) |
| Partially vaccinated | 19 (14.2%) | 38 (15.2%) |
| Fully vaccinated | 30 (22.4%) | 58 (23.2%) |
| **Baseline and end-line COVID-19 Rapid Antibody Test results** |  |  |
| Tested at baseline | 134 | 250 |
| Seropositive at baseline | 53 (39.6%) | 87 (34.8%) |
| Seronegative at baseline | 81 (60.4%) | 163 (65.2%) |
| No second test at end of follow-up <sup>a</sup> | 3 (3.7%) | 18 (11.0%) |
| Paired baseline seronegative samples tested for antibody at end of follow-up <sup>a</sup> | 78 (96.3%) | 145 (89.0%) |
| Seropositive at end of follow-up <sup>b</sup> | 17 (12.7%) | 39 (15.6%) |
| Seronegative at end of follow-up <sup>b</sup> | 61 (45.5%) | 106 (42.4%) |
| **Time to symptomatic HHC diagnosis from index case diagnosis** |  |  |
| n, | 1 | 12 |
| Mean days (SD) | 16.0 (0) | 15.9 (19.6) |

a. Denominator is those with seronegative test result at baseline
b. Denominator is those who had paired baseline and end of follow-up samples

In general, the symptomatic SARS-CoV-2 infection rate was markedly lower in the SCB-2019 recipient index case households and among their HHC compared with the infection rate in households and HHC of placebo recipient index cases (**Tables 3 and 4**).

**Table 3.** Symptomatic SARS-CoV-2 infection secondary attack rates (SAR) in the **households** of index cases.

| Households of SPECTRA index cases |  |  |  |  |  |  |  |  |  |
| --- | --- | --- | --- | --- | --- | --- | --- | --- | --- |
|  | SCB-2019 index cases (N = 51) |  |  |  | Placebo index cases (N = 103) |  |  |  | RRR <sup>c</sup><br>(90% CI*) |
|  | Cases | Households | Households with ≥ 1 case | Household SAR,<br>% of index case's contacts<br>(90% CI) <sup>a</sup> | Cases | Households | Households with ≥ 1 case | Household SAR,<br>% of index case's contacts<br>(90% CI) <sup>a</sup> |  |
| All | 1 | 51 | 1 | **0.76**<br>(0.15 –3.90) | 12 | 103 | 7 | **5.88**<br>(3.20–10.82) | **0.79**<br>(-0.28–0.97) |
| *Household COVID-19 vaccine coverage (%) <sup>b</sup>* |  |  |  |  |  |  |  |  |  |
| 75 to 100 | 0 | 9 | 0 | - | 2 | 25 | 2 | **4.27**<br>(1.33–13.73) | - |
| 50 to 74 | 0 | 12 | 0 | - | 0 | 18 | 0 | - | - |
| 25 to 49 | 0 | 7 | 0 | - | 8 | 16 | 3 | **18.3**<br>(7.48 –44.95) | - |
| 0 to 24 | 1 | 23 | 1 | **1.95**<br>(0.38 –9.87) | 2 | 44 | 2 | **2.48**<br>(0.77–8.00) | **0.21**<br>(-5.14–0.90) |

a. Point estimates and 90% CI for household secondary attack rates (**SAR**) and the indirect effect were estimated using a GEE logistic regression modeling approach with jack-knifed standard errors (see Methods).
b. Coverage for any COVID-19 vaccine received estimated as the proportion of household members who received any SARS-CoV-2 vaccine over total size of household.
c. Relative Risk Reduction (**RRR**) calculated as 1 minus the relative risk (**RR**) comparing SAR in SCB-2019 and Placebo households.

**Table 4.** Symptomatic SARS-CoV-2 infection secondary attack rates (SAR) among **household contacts** of index cases

|  | Household contacts of index cases |  |  |  | RRR <sup>a</sup> (90% CI<sup>b</sup>) |
| --- | --- | --- | --- | --- | --- |
|  | Cases among contacts /<br>Total number of contacts | SAR per 1000 persons<br>(90% CI) | Cases among contacts /<br>Total number of contacts | SAR per 1000 persons<br>(90% CI) |  |
| Secondary attack rates among household contacts of All Index Cases |  |  |  |  |  |
|  | SCB-2019 vaccine group (N = 134) |  | Placebo group (N = 250) |  |  |
| All Household Contacts | 1 / 134 | 7.5 (0.4–34.9) | 12 / 250 | 48.0 (27.9–76.6) | 0.84 (0.28–0.97) |
| Fully vaccinated | 0 / 30 | 0 (0–95.0) | 3 / 58 | 51.7 (14.2–128) | 1.00 (-0.66–1.00) |
| Partially vaccinated | 0 / 19 | 0 (0–146) | 1 / 38 | 26.3 (1.3–119) | 1.00 (-4.21–1.00) |
| Unvaccinated | 1 / 85 | 11.8 (0.6–54.6) | 8 / 154 | 51.9 (26.1–91.8) | 0.77 (-0.07–0.95) |
| Secondary attack rates among household contacts of Symptomatic Index Cases |  |  |  |  |  |
|  | SCB-2019 vaccine group (n = 106) |  | Placebo group (n = 212) |  |  |
| All Household Contacts | 1 / 106 | 9.4 (0.5–44.0) | 10 / 212 | 47.2 (25.8–78.7) | 0.80 (0.07–0.96) |
| Fully vaccinated | 0 / 21 | 0 (0–133) | 3 / 53 | 56.6 (15.6–140) | 1.00 (-1.13–1.00) |
| Partially vaccinated | 0 / 16 | 0 (0–171) | 1 / 33 | 30.3 (1.6–136) | 1.00 (-4.34–1.00) |
| Unvaccinated | 1 / 69 | 14.5 (0.7–66.9) | 6 / 126 | 47.6 (20.9–91.8) | 0.70 (-0.47–0.94) |
| Secondary attack rates among household contacts of Asymptomatic Index Cases |  |  |  |  |  |
|  | SCB-2019 vaccine group (n = 28) |  | Placebo group (n = 38) |  |  |
| All households | 0 / 28 | 0 (0–102) | 2 / 38 | 52.6 (9.4–157) | 1.00 (-0.76–1.00) |
| Fully vaccinated | 0 / 9 | 0 (0–283) | 0 / 5 | 0 (0–451) | - |
| Partially vaccinated | 0 / 3 | 0 (0–632) | 0 / 5 | 0 (0–451) | - |
| Unvaccinated | 0 / 16 | 0 (0–171) | 2 / 28 | 71.4 (12.8–208) | 1.00 (-1.21–1.00)- |

**SAR: Secondary Attack Rates; RRR: Relative Risk Reduction**
**a:** One minus the relative risk comparing the SAR among the HH contacts of SCB-2019 versus Placebo index cases
**b:** 90% confidence intervals estimated as 1 minus the score confidence interval for the ratio of two proportions [relative risk], as implemented in the *PropCIs* package (version 0.3.0) of R (version 4.1.2).

### Risk reductions in households

In the 51 households with an SCB-2019 vaccinee index case there was one household with a secondary infection (**Table 3**), an SAR of 76 (90% CI: 15–390), compared 7 of 103 households of placebo index cases with secondary infections for an SAR of 588 (320–1082); a relative risk reduction of 79% (90% CI: -28–97).

### Risk reductions in household contacts

One symptomatic secondary COVID-19 case among 134 HHC in the SCB-2019 index cases gave an SAR of 7.5 per 1,000 persons (90% CI: 0.4–34.9) compared with 12 symptomatic cases reported from 250 HHC in the placebo group, an SAR of 48.0 per 1,000 persons (27.9–76.6); a RRR of 84% (90% CI: 28–97) (**Table 4**). Risk reduction was similar across vaccinated and unvaccinated HHC who were distributed comparably between the two treatment groups. There were no cases of symptomatic SARS-CoV-2 infection in fully or partially vaccinated HHC of SCB-2019 recipient index cases. The RRR in symptomatic COVID-19 disease among unvaccinated HHC of SCB-2019 recipient index cases compared with unvaccinated HHCs of placebo recipient index cases was 77% (90% CI: -7–95).

When assessed for only those index cases with symptomatic COVID-19 the pattern was similar (**Table 4**). The SAR in the HHC of SCB-2019 index cases was 9.4 per 1000 person (90% CI: 0.5–44.0) compared with 47.2 per 1,000 persons (90% CI: 25.8–78.7) in placebo case HHC, a RRR of 80.0% (90% CI: 7–96). There were no secondary infections observed in 28 HHC of SCB-2019 index cases with asymptomatic COVID-19 infections, and only two cases in 38 HHC placebo index cases who had asymptomatic infections (**Table 4**). These small numbers preclude any meaningful analysis.

SAR for asymptomatic infections in HHC who were seronegative at baseline did not show any differences between those in households with SCB-2019 or placebo index cases with a RRR of 12% (90% CI: -32–43) based on SARs of 210 (90% CI: 138–298) and 239 (90% CI: 185–301) per 1,000 persons in SCB-2109 and placebo group households, respectively.

## DISCUSSION

Measures such as mask-wearing, social distancing, and work-from-home policies are major components of preventative health measures in the COVID-19 pandemic, but domestic transmission within households cannot be subject to such regulations. Therefore, a better understanding of this aspect of viral dissemination, and especially of how it is impacted by vaccination campaigns, is particularly important for health authorities to control outbreaks.

Our study findings, conducted as part of the SPECTRA trial with blinded vaccine and placebo groups, are an important contribution to understanding the impact of vaccination in preventing transmission among household contacts. The efficacy of SCB-2019 in the Philippines as part of the SPECTRA study was 69.3% (95% CI: 51.0–81.4) in the initial analysis of cases occurring a median of 24 days after the second dose; efficacy at 6 months post-vaccination was 48.3% (38.0–57.0) when most cases would have been Delta variant. Having monitored the occurrence of rRT-PCR-confirmed COVID-19 in household members of infected SPECTRA trial participants for three weeks, we found a 79% (90% CI: -28–97) risk reduction of secondary symptomatic infection rates in households if the COVID-19 positive index cases were vaccinated with SCB-2019 vaccine versus households where the infected index cases had received placebo in the parent SPECTRA trial. The 84% (90% CI: 28–97) reduction in secondary infections in household contacts of vaccinated index cases compared with placebo mirrored the trend seen in households. These results are particularly notable in being obtained when the Delta (B.1.617.2) variant predominated, a variant with more rapid symptom onset and higher viral load than earlier strains [22].

The study was conducted as part of a well-conducted, randomized, double-blind, placebo-controlled efficacy trial “SPECTRA” with a study team blinded to participant allocation throughout the study, providing a unique opportunity to minimize confounding while estimating the vaccine impact in preventing secondary infections among household contacts. Trial participants underwent intensive follow-up and testing for SARS-CoV-2 infection and disease. Any confirmed COVID-19 in SPECTRA participants in the Philippines was a potential index case in a household. Daily clinical follow-up of index case HHC used methods established for the clinical efficacy trial including weekly telephone call interviews and daily self-reported symptom checklists assessing HHC with signs and symptoms of any severity. All symptomatic cases were confirmed using rRT-PCR testing of NP samples following the WHO standard guidelines to ensure a reliable diagnosis of COVID-19 [20]. There were no cases of symptomatic SARS-CoV-2 infection in fully or partially vaccinated contacts of SCB-2019 recipient index case, indicating that the adjuvanted recombinant protein SCB-2019 vaccine provides both direct protection to the vaccinee and effectively exerts indirect effects preventing transmission. Whilst we were not able quantify it vaccinated people with breakthrough infections have been reported to have a faster decline in viral load than unvaccinated persons [18], which significantly impacts transmission [23].

Household transmission has played a significant role in the scale of the COVID-19 pandemic [21,24]. There is a lack of conclusive evidence on the impact of vaccines to prevent secondary infections in household contacts; some studies have demonstrated a positive impact of COVID-19 vaccines on reducing household transmission of SARS-CoV-2 [12–16] while others reported limited impact in decreasing the risk of household transmission of COVID-19 especially for contacts of Delta variant index cases [17,18]. A major barrier to obtaining conclusive evidence is the lack of control groups and the presence of significant confounding, so our study findings, with blinded vaccine and placebo groups, provides important information on the impact of vaccination on secondary COVID-19 infections among HHC, and particularly on the impact of SCB-2019 on preventing such secondary infections in HHC.

Elucidating the role of household transmission can have several policy and practice implications particularly to inform vaccination practices and policies in resource-limited countries where the coverage is still low such as sub–Saharan Africa where average vaccine coverage is approximately 10% [25]. In countries with limited access to COVID-19 vaccines a focus on households could support optimal vaccine use to control of SARS-CoV-2 transmission. As COVID-19 can transition to endemicity in many settings, understanding the mechanisms and developing prevention strategies for household transmission will provide several opportunities. Moreover, with the potential for future waves as vaccine immunity wanes or new variants emerge, vaccination strategies globally will need to consider household coverage of COVID-19 vaccination as an important aspect of limiting viral transmission.

### Limitations

Our study findings should be interpreted with several important limitations. First, many index cases refused consent for this household study due to fear of stigma and discrimination related to COVID-19 infection in the Philippines which could have created a selection bias if different between SCB-2019 and placebo index cases. However, of the 441 refusals 155 (35%) and 286 (65%) had received SCB-2019 or placebo, respectively, mirroring the 51 (33%) and 103 (67%) who did enroll. Secondly, detection of incident asymptomatic infections during the follow-up of HHC using RAT kits may underestimate the burden of asymptomatic COVID-19 infections due to their potentially limited sensitivity and specificity [26–28]. Third, pandemic-related lockdowns and travel restrictions disrupted onsite monitoring and quality control/assurance activities, but data quality was assured through centralized database-based monitoring and contracting local monitors for on-site visits. Crude secondary attack rate estimates presented here do not account for possible exposure to sources of SARS-CoV-2 infection other than the SPECTRA trial index case, an issue in practically all household contact studies. The lack of multiple variable adjustment makes it likely that these SAR estimates are affected by residual confounding. Finally, we do not know whether transmission of more infectious Omicron variants would be affected, although experience suggests that impact on severe outcomes would be retained.

## CONCLUSION

Based on a limited number of households and contacts we observed a trend of 79% reduction in household transmission and 80% in HHC transmission for SCB-2019 vaccinated index cases rather than placebo in an environment of predominantly Delta variant circulation.

## Data Availability

All data produced in the present work are contained in the manuscript

## FUNDING

This work was supported by the Coalition for Epidemic Preparedness Innovations (Grants RRCL 2001 and RCL2202) and Clover Biopharmaceuticals.

## ACKNOWLEDGEMENTS

The authors are grateful to all participants in the study and to our external service providers (CROs, laboratories, clinical suppliers and biostatistics) for their invaluable assistance in performing the trial. We thank Keith Veitch (keithveitch communications, The Netherlands) for editorial assistance with the manuscript (funded by Clover Biopharmaceuticals).

## POTENTIAL CONFLICTS OF INTEREST

P.L. and J.G. are full-time employees of the study sponsor. Other authors declare they have no conflicts.

## Supplementary material

**Supplementary table 1:** Study sites and enrolled index cases in each site

| **Study Site** | **Number of Index cases (%)** | **Number of HH contacts (%)** |
| --- | --- | --- |
| University of Philippines, Manila | 27 (17.2) | 62 (15.9) |
| Asian Hospital and Medical Center | 35 (22.3) | 73 (18.7) |
| Tropical Disease Foundation | 22 (14.0) | 36 (9.2) |
| Manila Doctors Hospital | 20 (12.7) | 70 (17.9) |
| Las Piñas Doctors Hospital | 21 (13.4) | 58 (14.8) |
| UERM Memorial Medical Center-Laguna | 2 (1.3) | 2 (0.5) |
| FEU-NRMF Clinical Research | 28 (17.8) | 85 (21.7) |
| UERM Memorial Medical Center-Quezon City | 2 (1.3) | 5 (1.3) |
| **Total** | **157** | **391** |

**Supplementary table 2.** Medical histories and vaccination records of household contacts

|  | Household contacts of index cases |  |  |
| --- | --- | --- | --- |
|  | Total | SCB-2019 group | Placebo group |
|  | (N = 384) | (n = 134) | (n = 250) |
| Any underlying conditions | 36<br>(9.4%) | 9<br>(6.7%) | 27<br>(10.8%) |
| Pregnancy | 3<br>(0.8%) | 1<br>(0.7 %) | 2<br>(0.8%) |
| Cardiovascular disease (including hypertension) | 14<br>(3.6%) | 4<br>(3.0%) | 10<br>(4.0%) |
| Diabetes | 3<br>(0.8%) | 0<br>(0%) | 3<br>(1.2%) |
| Chronic neurological disease | 1<br>(0.3%) | 0<br>(0%) | 1<br>(0.4%) |
| Renal or hepatic disease, or HIV | 0<br>(0%) | 0<br>(0%) | 0<br>(0%) |
| Other | 18<br>(4.7%) | 4<br>(3.0%) | 14<br>(5.6%) |
| Vaccination history |  |  |  |
| Fully vaccinated | 88<br>(22.9%) | 30<br>(22.4%) | 58<br>(23.2%) |
| Partially vaccinated | 57<br>(14.8%) | 19<br>(14.2%) | 38<br>(15.2%) |
| Not vaccinated | 239<br>(62.2%) | 85<br>(63.4%) | 154<br>(61.6%) |

There were no significant differences between SCB-2019 and placebo groups when tested by Pearson's Chi-Squared Test or Fisher's Exact Test

## Notes

### Author Declarations

The protocol was approved by the Single Joint Research Ethics Board (SJREB), the respective Institutional Review Boards (IRB) of the eight study sites and the International Vaccine Institute IRB

